# Reference values of Complete blood count in healthy adult Eritrean

**DOI:** 10.1101/2023.03.02.23286685

**Authors:** Ahmed O Noury, Omer A. Musa, Elmuiz Gasmalbari, Barakat M Bakhit, Eyasu H Tesfamariam, Daniel M Abraha, Zekarias B Ghebre, Omer Suleman, Efrem G Tesfay, Filmon G Hailezghi

## Abstract

**Background:** Blood count is the most commonly prescribed biological examination in general medical practice. The reference intervals of the hematological parameters of this examination are critical for clinical orientations and therapeutic decisions. Because there are racial, ethnic, and geographical differences in complete blood count (CBC) reference intervals (RIs), population-specific RIs must be established. The goals of this study were to identify hematological reference ranges in healthy adult Eritreans.

**Method:** 942 healthy Eritreans between 18 and 60 years old were included,331 males and 611 females by use of a DXH500 analyzer, age ranging between 18-60 years. The venous blood sample was collected in a tube containing EDTA anticoagulant for the blood tests. SPSS version 25 statistical software was used for data analysis, P value < 0.05 was considered significant A non-parametric test was used for the determination of upper (97.5th percentile) and lower (2.5th percentile) reference interval limits with 95% CI. The Harris and Boyd Rule is used to determine the need for partitioning of reference intervals based on gender.

**Results:** The established 95% reference intervals combined median (2.5th–97.5th percentile) for both males and females were: WBCs: 6.37 (3.02-13.55×103/µL), Lymph%:39.34 (21.39-60.54 %), Mono %:8.98 (5.18-14.54%), Neut%: 49.13(16.90-81.98 %), Baso%: 0.22 (0.00-0.63%), MCV: 87.67 (76.58-97.29fl), MCH: 27.53 (20.46-32.70 pg), MCHC: 31.38 (25.20-35.30 g/dl, RDW: 14.65 (12.70-18.60 %), PLT: 286.83 (131.62-453.13 ×103/µL) and MPV: 8.92 (7.28-11.01fl). The parameters that demand separate RI and Their respective median (2.5th – 97.5th percentile) for males versus females were: Eosin: 3.86 (0.29-16.68 %) versus 1.80 (0.20-6.73 %), RBCs: 5.57 (4.47-7.69×106/µL) versus 4.97 (3.98-6.38×106/µL), Hb: 15.28 (11.48-17.99 g/dl) versus 13.50 (10.74-16.54 g/dl), and HCT: 48.75 (38.96-61.17 %) versus 43.19 (34.86-58.60 %).

The median of WBCs was significantly higher in females than males, the mean WBCs were lower in people residing at high altitudes compared to those leaving at low altitudes, The WBC is significantly higher among obese participants. The median Platelet count is significantly higher in females than in males.

**Conclusion:** The reference intervals established in this study differ from the international one and thus should be used for the interpretation of laboratory results in diagnosis and follow-up in Eritrea. The study showed significant variations in Hb levels, RBCs count, WBCs count, and platelet according to gender, Age, BMI, and physical activity.

## Introduction

Complete cell count (CBC) is the most frequently used laboratory investigation in health care. The most important aspect of laboratory test interpretation is the concept of reference interval (RI), where test values that fall inside the range are considered normal and those occurring outside the range are considered abnormal^(1)^. Population-based RI, which first came into use in human medicine in 1969, is the range of values that 95% of a healthy population fell within ^(2)^. The reference ranges universally used are mostly from studies conducted in Western countries. However, these reference values differ based on many socio-demographic characteristics. Several factors such as age, gender, dietary pat-terns, ethnic differences and altitude affect the reference ranges for different groups ^(3)^and the Western RI may not be applicable in most regional settings. The Clinical and Laboratory Standards Institute (CLSI) recommended each country should establish its organic standards to avoid misinterpretation of blood count results^(4)^. The objective of this study is to determine the Hematological reference values in healthy adult Eritreans

## 2. Materials and Methods

### 2.1 Study design

This is descriptive and cross-sectional

### 2.2 Study site

The study was conducted on Eritreans from all over the country in the period from 2019 up to 2022. Samples used in this study were collected from the following sites in Eretria:

— Central region (Asmara)
— Anseba region (Keren)
— Southern region (Mendefera)
— Northern Red Sea region (Massawa)

### 2.3 Volunteer recruitment

The areas from which participants were drawn were chosen using a multistage sampling technique (Stratified sample). The zone was the first-stage sampling unit. Out of several zones in the city, we chose one at random. The second stage involved a random sub-zone selection. We chose one of the zone’s sub-zones at random. The third stage was the random selection of sub-zone blocks. We chose four blocks at random from the sub-zone. The fourth stage involved systematically selecting households from a randomly selected reference point. After identifying a household, we contacted the zone administrators and requested that one employer be assigned to educate the population of the intended blocks about the study’s objectives. Recruitment was stratified into 4 age groups: 18-29, 30-39, 40-49, and 50-60 years.

### 2.4 Reference population

This cross-sectional study included 942 healthy adult Eritreans from various social, ethnic, and professional groups. The Clinical and Laboratory Standards Institute guidelines were used to select participants for this study. Our reference sample included 331 men with an average age of 40 and 611 women with an average age of 41; the study lasted from November 2019 to January 2022.

### 2.5 Ethical clearance

The Eritrean Ministry of Health’s ethics committee granted ethical approval for this study. Before drawing blood, we obtained written informed consent from all participants. The team used a questionnaire to collect anthropometric measurements, demographic data, medical status, medical history, physical activity, and sleeping hours from each blood donor. Blood pressure and BMI measurements were taken for all participants.

### 2.6 Blood Collection

An early morning visit was from 8 AM to 12 AM. Blood samples were collected by a trained phlebotomist, blood drawn by venipuncture 2 ml of venous blood was collected into tetra-acetic acid (EDTA) vacutainer tubes for a complete blood count. Blood samples were transported to the laboratory, where all the analyses were conducted at the Hematology Department within 5 hours of blood extraction.

### 2.7 Statistical Analysis

Analysis was done in SPSS (Version 25) after a careful checkup on completeness, cleaning, and editing processes. Extreme values that might greatly affect the result within each gender were identified using the D/R ratio, where D is the absolute difference between an extreme observation (large or small) and the next largest (or smallest) observation, and R is the range (maximum-minimum). The identified extreme values were deleted if D/R≥1/3 (4). Descriptive analysis of the demographic variables, stratified by gender, was performed using mean, median, standard deviation, frequencies, and percent, as appropriate. The Clinical Laboratory Standards Institute /International Federation for Clinical Chemistry (CLSI/IFCC) was employed to compute the reference intervals. As per the standard, a nonparametric method median (IQR), range (Minimum and maximum), combined and separate 95% RIs (2.5th and 97.5th percentiles), 95% CI for the lower limit of RI, and upper limit of RI were computed (using 1000 bootstrapped simple random sampling). Harris and Boyd test vis-à-vis Mann-Whitney U test, was performed to determine whether combined or separate RIs are needed (5,6). However, results from Harris and Boyd were finally endorsed. Upon using Harris and Boyd, the statistical Z result was compared with a critical Z* value: Where 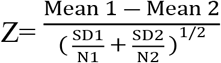 and Z*=3[(N1+N2)/240]1/2.

Separate gender specific reference range are needed when Z>Z*.

To assess the relationship of the hematological parameters with the socio-demographic and basic background characteristics, the spearman rank correlation (for continuous variables), Mann-Whitney u test (categorical variables having dichotomous outcome), and Kruskal-Wallis test (categorical variables having more than two outcomes) were employed. Post-hoc analysis using the Bonferroni test was also performed for the results that were found to be significant using Kruskal-Wallis. Agreement between the currently estimated reference intervals and currently used RIs was performed using the Kappa statistic. Interpretation of the Kappa statistic was done using Landis and Koch classification (7).

## 3. Results

A total of 942 individuals volunteered to participate in our study and were included, including 331 males and 611 females. The percentages of male and female participants were 35.14% and 64.86% respectively. Combined and separate (male and female) descriptive characteristics, which include mean (SD), median (IQR), and range, of study participants for the four cities are displayed in Table 1. The combined mean age was 39.69 (±12.38) years. The combined mean (SD) of height and weight were 1.62 (±0.09) meters and 58.85 (12.23) kilograms, respectively. The combined mean body mass index was also 22.49 (±4.56) kg/m2. Moreover, the combined mean (SD) values of SBP and DBP were 118.03 (16.32) and 76.32 (9.69) respectively. The mean sleeping hour (per day) of both males and females was 7.75 (0.86) hours. A detailed characterization of the study participants stratified by gender is given in Table 1 (continuous variables) and Table 2 (categorical variables).

**Table 1:** Socio-demographic and basic background characteristics of the study participants for continuous variables stratified by gender

| Continuous variable | Combined | Male | Female |
| --- | --- | --- | --- |
| **Age (Years)** |  |  |  |
| Mean (SD) | 39.69 (12.38) | 37.00 (13.24) | 41.16 (11.58) |
| Median (IQR) | 40.00 (21) | 34.50 (25) | 42.00 (19) |
| **Height (m)** |  |  |  |
| Mean (SD) | 1.62 (0.09) | 1.70 (0.07) | 1.57 (0.06) |
| Median (IQR) | 1.61 (0.13) | 1.7 (0.10) | 1.57 (0.09) |
| **Weight (kg)** |  |  |  |
| Mean (SD) | 58.85 (12.23) | 61.81 (12.09) | 57.24 (12.01) |
| Median (IQR) | 57.9 (16) | 60.10 (15) | 56 (16) |
| **Body Mass index (kg/m<sup>2</sup>)** |  |  |  |
| Mean (SD) | 22.49 (4.56) | 21.36 (4.12) | 23.1 (4.65) |
| Median (IQR) | 22.03 (6.2) | 21.02 (5) | 22.72 (6.7) |
| **SBP** |  |  |  |
| Mean (SD) | 118.03 (16.32) | 117.86 (14.24) | 118.18 (17.37) |
| Median (IQR) | 118 (17) | 120 (16) | 118 (20) |
| **DBP** |  |  |  |
| Mean (SD) | 76.32 (9.69) | 76.8 (8.55) | 76.03 (10.23) |
| Median (IQR) | 78 (10) | 78 (10) | 78 (10) |
| **Sleeping Hours (per day)** |  |  |  |
| Mean (SD) | 7.75 (0.86) | 7.68 (0.81) | 7.78 (0.88) |
| Median (IQR) | 8 (0.0) | 8 (0.0) | 8 (0.0) |

**SD: Standard deviation, IQR: Interquartile range**

**Table 2:** Socio-demographic and basic background characteristics of the study participants for categorical variables stratified by gender

| Categorical Variable | Combined |  | Male |  | Female |  |
| --- | --- | --- | --- | --- | --- | --- |
|  | N | % | N | % | N | % |
| **City** |  |  |  |  |  |  |
| Asmara | 401 | 42.4 | 128 | 38.7 | 273 | 44.7 |
| Massawa | 163 | 17.3 | 74 | 22.4 | 87 | 14.2 |
| Mendefera | 177 | 18.7 | 50 | 15 | 127 | 20.8 |
| Keren | 204 | 21.6 | 79 | 23.9 | 124 | 20.3 |
| **Education** |  |  |  |  |  |  |
| Illiterate | 153 | 16 | 26 | 7.3 | 127 | 20.8 |
| Primary | 137 | 14.5 | 28 | 8.5 | 108 | 17.7 |
| Junior | 252 | 26.7 | 66 | 19.9 | 186 | 30.4 |
| Secondary | 277 | 29.3 | 120 | 36.3 | 157 | 25.7 |
| College | 123 | 13 | 92 | 27.8 | 31 | 5.1 |
| **Marital Status** |  |  |  |  |  |  |
| Married | 659 | 69.7 | 197 | 59.5 | 461 | 75.5 |
| Widowed | 24 | 2.5 | 1 | 0.3 | 23 | 3.8 |
| Divorced | 25 | 2.6 | 1 | 0.3 | 24 | 3.9 |
| Separated | 5 | 0.5 | 0 | 0 | 5 | 0.8 |
| Single/Never married | 226 | 23.9 | 131 | 39.6 | 95 | 15.5 |
| **Ethnic Group** |  |  |  |  |  |  |
| Afar | 27 | 2.9 | 13 | 3.9 | 14 | 2.3 |
| Blen | 87 | 9.2 | 38 | 11.5 | 49 | 8 |
| Nara | 1 | 0.1 | 1 | 0.3 |  |  |
| Saho | 29 | 3.1 | 8 | 2.4 | 20 | 3.3 |
| Tigre | 142 | 15 | 76 | 23 | 66 | 10.8 |
| Tigrigna | 656 | 69.4 | 195 | 58.9 | 461 | 75.5 |
| **Exercise** |  |  |  |  |  |  |
| Yes | 172 | 18.3 | 119 | 36.1 | 53 | 8.7 |
| No | 770 | 81.7 | 211 | 63.9 | 558 | 91.3 |
| **BMI** |  |  |  |  |  |  |
| Underweight | 141 | 14.9 | 59 | 17.8 | 82 | 13.4 |
| Normal weight | 534 | 56.5 | 209 | 63.1 | 324 | 53 |
| Overweight | 207 | 21.9 | 53 | 16 | 154 | 25.2 |
| Obese | 63 | 6.7 | 10 | 3 | 51 | 8.3 |
| **Sleeping Hours (per day)** |  |  |  |  |  |  |
| Less than 6 hours | 25 | 2.6 | 10 | 3 | 15 | 2.5 |
| 6-8 hours | 849 | 89.9 | 301 | 90.9 | 547 | 89.5 |
| More than 8 hours | 62 | 6.6 | 13 | 3.9 | 49 | 8 |

Others\* include Afar, Saho, and Blen.

### Parameters that demand partitioned RI and their distribution

As per the Harris and Boyd recommendation for the need in partitioning the reference interval by gender four hematological parameters, namely, Eosin, RBC, Hb, and HCT were essentially found to have separate RI (Table 3).

**Table3.** Hematological parameters and need for partitioning of reference intervals by gender.

| Parameters | Harris and Boyd |  | Decision |
| --- | --- | --- | --- |
|  | Z | Z* |  |
| WBCs 103/μL | 1.59 | 5.94 | Combined |
| Lymphocytes (%) | 2.26 | 5.94 | Combined |
| Monocytes (%) | 2.04 | 5.94 | Combined |
| Neutrophils (%) | 0.12 | 5.94 | Combined |
| Eosinophils (%) | 8.02 | 5.94 | Separate |
| Basophils (%) | 3.48 | 5.92 | Combined RI |
| RBCs $\times 106/\mu$L | 12.99 | 5.94 | Separate RI |
| HB (g/dl) | 17.92 | 5.94 | Separate RI |
| HCT (%) | 15.32 | 5.94 | Separate RI |
| MCV (fl) | 1.62 | 5.94 | Combined RI |
| MCH (pg) | 1.93 | 5.94 | Combined RI |
| MCHC (g/dl) | 1.46 | 5.94 | Combined RI |
| RDW (%) | 0.52 | 5.94 | Combined RI |
| RDW-SD | 0.64 | 5.94 | Combined RI |
| Platelets $\times 103/\mu$L | 2.87 | 5.93 | Combined RI |
| MPV (fl) | 5.21 | 5.93 | Combined RI |

Z\*, critical value, Z, calculated value, Separate RI are needed on when Z is greater than Z\*; $S_1^2$ is different from $S_2^2$ .

To make comparisons with other studies, the reference intervals for all the parameters were computed separately (males and females) as well as the combined data.

### Hematological Reference Intervals

Table 4 shows the mean (SD), median (IQR), range (minimum to maximum), 95% reference range (2.5th to 97.5th percentile), 95% CI for the lower limit (2.5th percentile), and 95% CI for the upper limit (97.5th percentile).

**Table 4:**
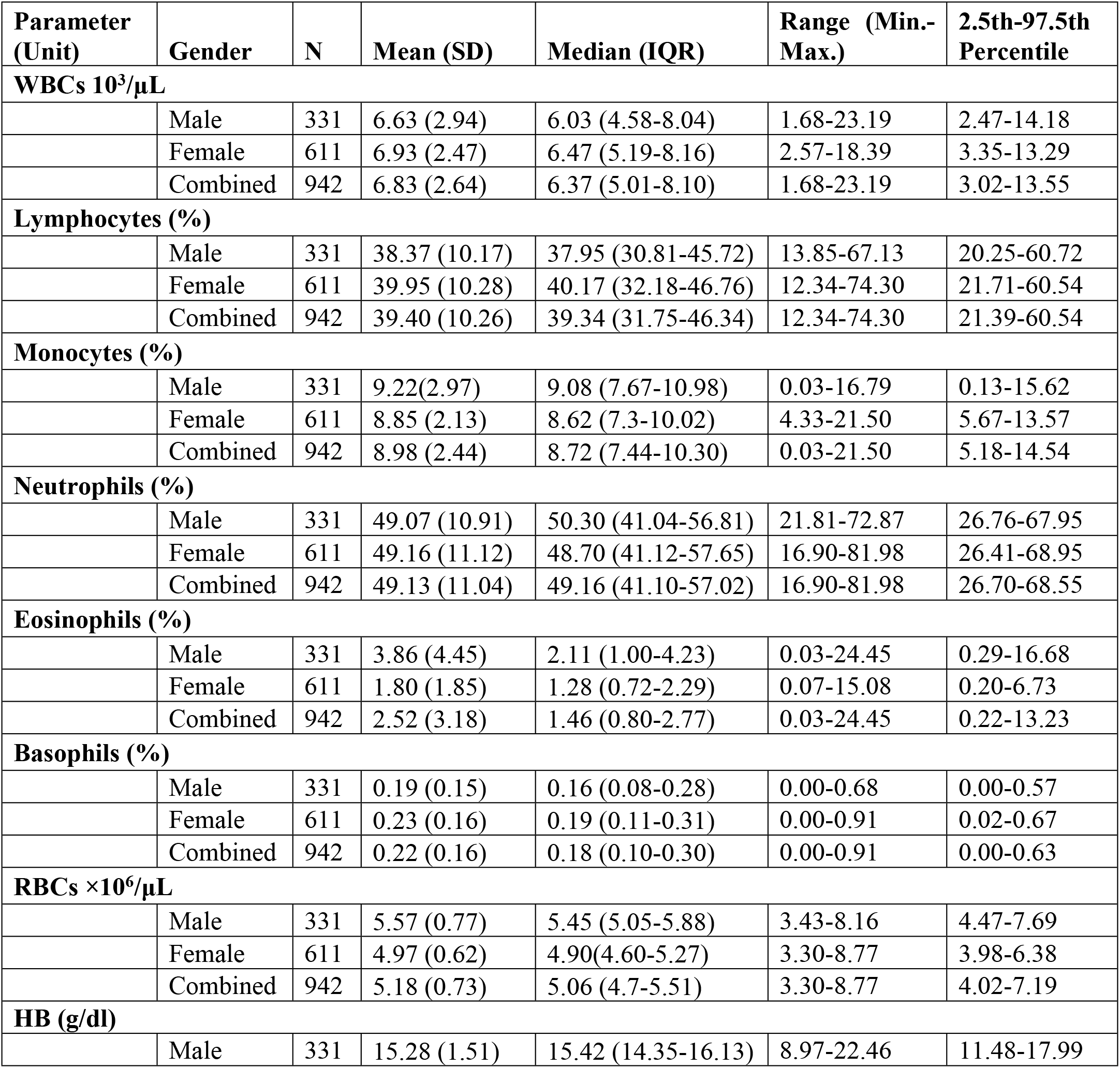
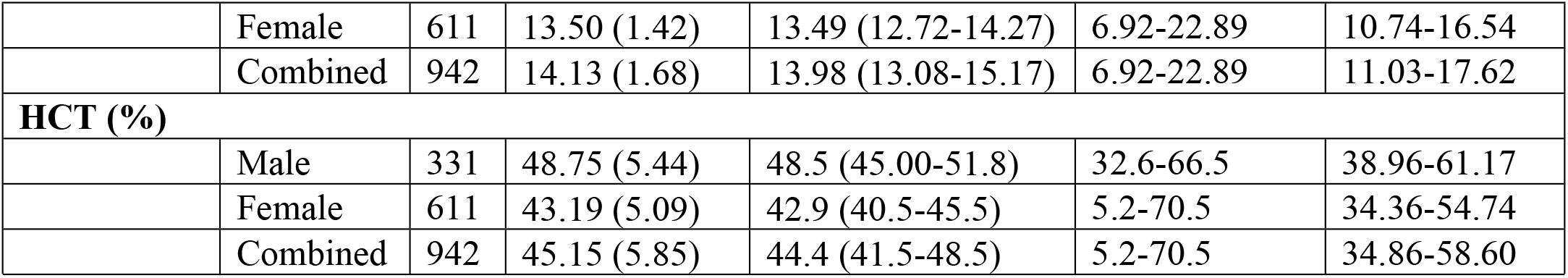
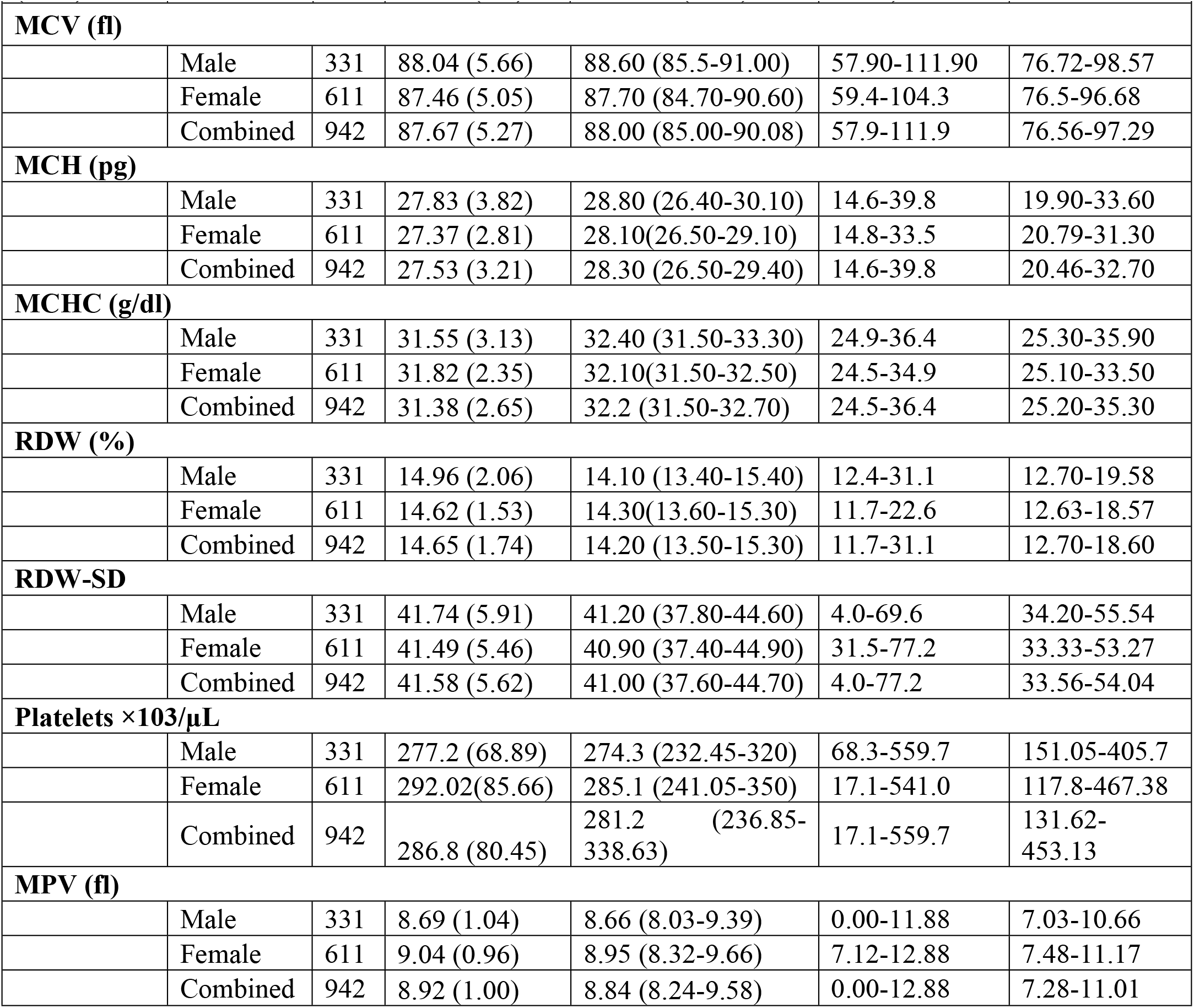
Complete descriptive analysis of the hematological parameters along with the 95% reference range (2.5^th^ to 97.5^th^ percentile)

**Table 5.** Spearman’s correlation of the hematological parameters with the continuous background.

| Parameters | Age | Height | Weight | BMI | SBP | DBP |
| --- | --- | --- | --- | --- | --- | --- |
|  | r (p-value) | r (p-value) | r (p-value) | r (p-value) | r (p-value) | r (p-value) |
| **WBCs 103/μL** | 0.049<br>(0.134) | -<br>0.108** (0.001) | 0.042<br>(0.202) | 0.118***<br>($<0.001$) | 0.087**<br>(0.008) | 0.071*<br>(0.030) |
| **Lymphocytes(%)** | 0.059<br>(0.069) | -0.078*<br>(0.017) | -0.013<br>(0.697) | 0.024<br>(0.458) | 0.072*<br>(0.029) | 0.117***<br>($<0.001$) |
| **Monocytes (%)** | 0.025<br>(0.448) | 0.812<br>(0.051) | -0.135***<br>($<0.001$) | -0.130***<br>($<0.001$) | 0.048<br>(0.144) | 0.047<br>(0.156) |
| **Neutrophils (%)** | -0.042<br>(0.196) | 0.051<br>(0.119) | 0.045<br>(0.164) | 0.029<br>(0.381) | -0.056<br>(0.087) | -0.101**<br>(0.002) |
| **Eosinophils (%)** | -0.124***<br>($<0.001$) | 0.170***<br>($<0.001$) | 0.031<br>(0.342) | -0.074*<br>(0.024) | -0.068*<br>(0.039) | -0.045<br>(0.168) |
| **Basophils (%)** | 0.149***<br>($<0.001$) | -0.105<br>(0.001) | -0.009<br>(0.792) | 0.054 (0.1) | 0.039<br>(0.232) | -0.019<br>(0.572) |
| **RBCs $\times 106/\mu$L** | -0.127***<br>($<0.001$) | 0.253***<br>($<0.001$) | 0.070*<br>(0.032) | -0.061<br>(0.062) | 0.031(0.339) | 0.074*<br>(0.025) |
| **HB (g/dl)** | -0.144***<br>($<0.001$) | 0.417***<br>($<0.001$) | 0.189***<br>($<0.001$) | -0.041<br>(0.214) | 0.022<br>(0.501) | 0.047<br>(0.151) |
| **HCT (%)** | -0.127***<br>($<0.001$) | 0.331***<br>($<0.001$) | 0.066*<br>(0.044) | -0.102**<br>(0.002) | 0.027<br>(0.404) | 0.048<br>(0.142) |
| **MCV (fl)** | 0.017<br>(0.603) | 0.077*<br>(0.018) | -0.012<br>(0.703) | -0.058<br>(0.077) | -0.028<br>(0.389) | -0.083*<br>(0.011) |
| **MCH (pg)** | -0.038<br>(0.239) | 0.172***<br>($<0.001$) | 0.078*<br>(0.017) | -0.025<br>(0.438) | -0.022<br>(0.498) | -0.047<br>(0.156) |
| **MCHC (g/dl)** | -0.075*<br>(0.021) | 0.208***<br>($<0.001$) | 0.116***<br>($<0.001$) | -0.007<br>(0.827) | -0.007<br>(0.828) | 0.01<br>(0.763) |
| **RDW (%)** | 0.047<br>(0.152) | -0.108**<br>(0.001) | -0.193***<br>($<0.001$) | -0.132***<br>($<0.001$) | 0.027<br>(0.408) | 0.059<br>(0.071) |
| **RDW-SD** | 0.04 (0.222) | -0.059<br>(0.07) | -0.228***<br>($<0.001$) | -0.197***<br>($<0.001$) | -0.009<br>(0.788) | -0.008<br>(0.818) |
| **Platelets $\times 103/\mu$L** | -0.096**<br>(0.003) | -0.082*<br>(0.012) | -0.006<br>(0.847) | 0.046<br>(0.158) | -0.03<br>(0.360) | -0.061<br>(0.063) |
| **MPV (fl)** | 0.093**<br>(0.004) | -0.102**<br>(0.002) | 0.080*<br>(0.015) | 0.145***<br>($<0.001$) | 0.038<br>(0.247) | 0.035<br>(0.289) |

\*p value $<0.05$ , \*\*p-value $<0.01$ , \*\*\*p-value $<0.001$

As per the recommendation of Harris and Boyd, the combined median (2.5^th^ – 97.5^th^percentile) for both males and females of WBC, Lym, Mono, Neut, Baso, MCV, MCH, MCHC, RDW, RDW-SD, PLT and MPV were 6.37 (3.02-13.55×103/µL), 39.34 (21.39-60.54 %), 8.98 (5.18-14.54%), 49.13(16.90-81.98 x%), 0.22 (0.00-0.63%), 87.67 (76.58-97.29fl),27.53 (20.46-32.70 pg), 31.38 (25.20-35.30 g/dl), 14.65 (12.70-18.60 %), 41.58 (33.56-54.04), 286.83 (131.62-453.13 ×10^3^/µL), and 8.92 (7.28-11.01fL) respectively.

The parameters that demand separate RI were Eosin, RBC, Hb, and HCT. Their respective median (2.5^th^ – 97.5^th^ percentile) for males versus females were 3.86 (0.29-16.68 %) versus 1.80 (0.20-6.73 %), 5.57 (4.47-7.69×10^6^/µL) versus 4.97 (3.98-6.38×10^6^/µL), 15.28 (11.48-17.99 g/dl) versus 13.50 (10.74-16.54 g/dl), and 48.75 (38.96-61.17 %) versus 43.19 (34.86-58.60%).

## DISCUSSION

The purpose of this study was to determine the hematological reference ranges of Eritrean healthy adults, identify factors that influence these values, and compare them to international and neighboring country values. In our study, Eritrean total white blood cell counts were similar to Caucasians, but we found an increased lymphocyte count and a decreased neutrophil count in comparison to Caucasians. (8,9). These findings are in contrast to the study carried out by Chen et al (10) who found that White subjects had a significantly higher WBC count in all age groups in comparison to black subjects. Many African Americans have WBCs that are persistently below the normal range for people of European descent, a condition called “benign ethnic Neutropenia. David et al explained that neutropenia in Africans is due to a regulatory variant in the Duffy antigen receptor for the chemokines gene (11). In our study, we found that Eritreans had higher hematocrit levels, higher hemoglobin levels, and higher RBCs than Caucasian and Africans. We hypothesized that the cause for this difference is likely due to altitude deference and Nutritional behavior; which is one of the factors affecting Hb and RBCs indices, Injera is the most important component of food in Eritrea. Teff is the main ingredient in injera, Teff is rich in carbohydrates, and fiber and has a complete set of essential amino acids. Teff is also particularly high in iron and has more calcium, copper, and zinc than other cereal grains (12). in this regard, Alaunyte(13) stated that The high Fe content of teff is reflected by the low prevalence of anemia in places where the teff grain is predominantly consumed

The study made by Mohammed (14) in pregnant Ethiopian women revealed that a decrease in the frequency of teff consumption was significantly associated with an increase in the likelihood of anemia We found significant gender effects on the CBC. Female participants had higher WBC, Lym, Baso, PLT, and MPV. while males had higher Mono, Eosin, RBC, HB, HCT, MCV, MCH,and MCHC The reasons for these differences have been attributed to factors such as the influence of the androgen hormone on erythropoiesis and decreased metabolic demand, decreased muscle mass, and lower iron stores due to menstruation in females, In this regard our findings are in agreement with previous studies(15–17)

In a recent study, Bachman et al (18)reported significantly increased erythropoietin levels and decreased ferritin and hepcidin with testosterone administration.

Interestingly, exogenous testosterone was used initially as a treatment for anemia. The American practice guideline on testosterone therapy recommends against the use of testosterone in patients with a hematocrit above 50% (19)

In support of our findings of significantly increased Monocytes in males, many studies showed a decrease in the number of Monocytes when estrogen level is high (20,21)

A new study by <u>Thiago</u> et al reveals testosterone administration in men increases Monocyte counts. (22)

Our results agree with those from a recent study by Hartlet al found that Eosinophil is higher in males than in females in all age groups (23)

In this study, the mean WBCs, MPV, PLT, and Baso were significantly higher for women than men.

Rukia et al(24)in their study found that the women had higher white blood cell and platelet counts compared to men, which agrees with our findings. Whereas Kaya et al.(25) Have found significantly higher WBCs in men compared to females which are inconsistent with our findings. The significant increase in platelet count and MPV in females might be due to the effect of estrogen on Megakaryopoiesis and platelet production according to previous research (26–28) Our study also indicated that the variation among the data sets for Eosin, Baso, hemoglobin, RBC, and HCT of the four age groups was statistically significant. In the present study we found that with an increase in age, there is a significant increase in Baso. The present results are consistent with previous studies (29,30). **O**ther reports show no age-related changes in Basophile count (31). The effects of aging on basophil must be explained by many mechanisms. First, aging increases basophil sensitivity to anti-IgE and IgE-mediated releaseability of histamine.(32)

Second, with aging, gut microbiota composition changes. (33) Basophil hematopoiesis and function are regulated by gut microbiota. The absence of gut microbiota leads to increased Basophil frequencies (34)

However, with an increase in age, a significant decrease in RBC, HB, HCT, and PLT was observed. Similar results have been reported by Isabel et al (35) in their study MCV, MCH, and RDW showed a significant positive correlation with age, hemoglobin, hematocrit, RBC, and platelet count showed a significant negative correlation. With the aging process the amount of growth hormone secreted declines, it is commonly believed that growth hormone stimulates erythropoiesis by increasing the oxygen consumption of tissues and thereby promoting tissue hypoxia, which in turn accelerates erythropoietin production by the kidneys. (36) Another hypothesis concerning the mechanisms that are responsible for the age-related changes of reduction may reflect a reduction in hematopoietic stem cell reserve during aging (37). Therefore, there is a definite need to introduce age-specific reference ranges.

## Data Availability

All relevant data are within the manuscript and its Supporting Information files.

